## Supplementary File for "Seasonal variations in social contact patterns in a rural population in north India: Implications for pandemic control"

### Supplementary Information

#### Supplementary Figures

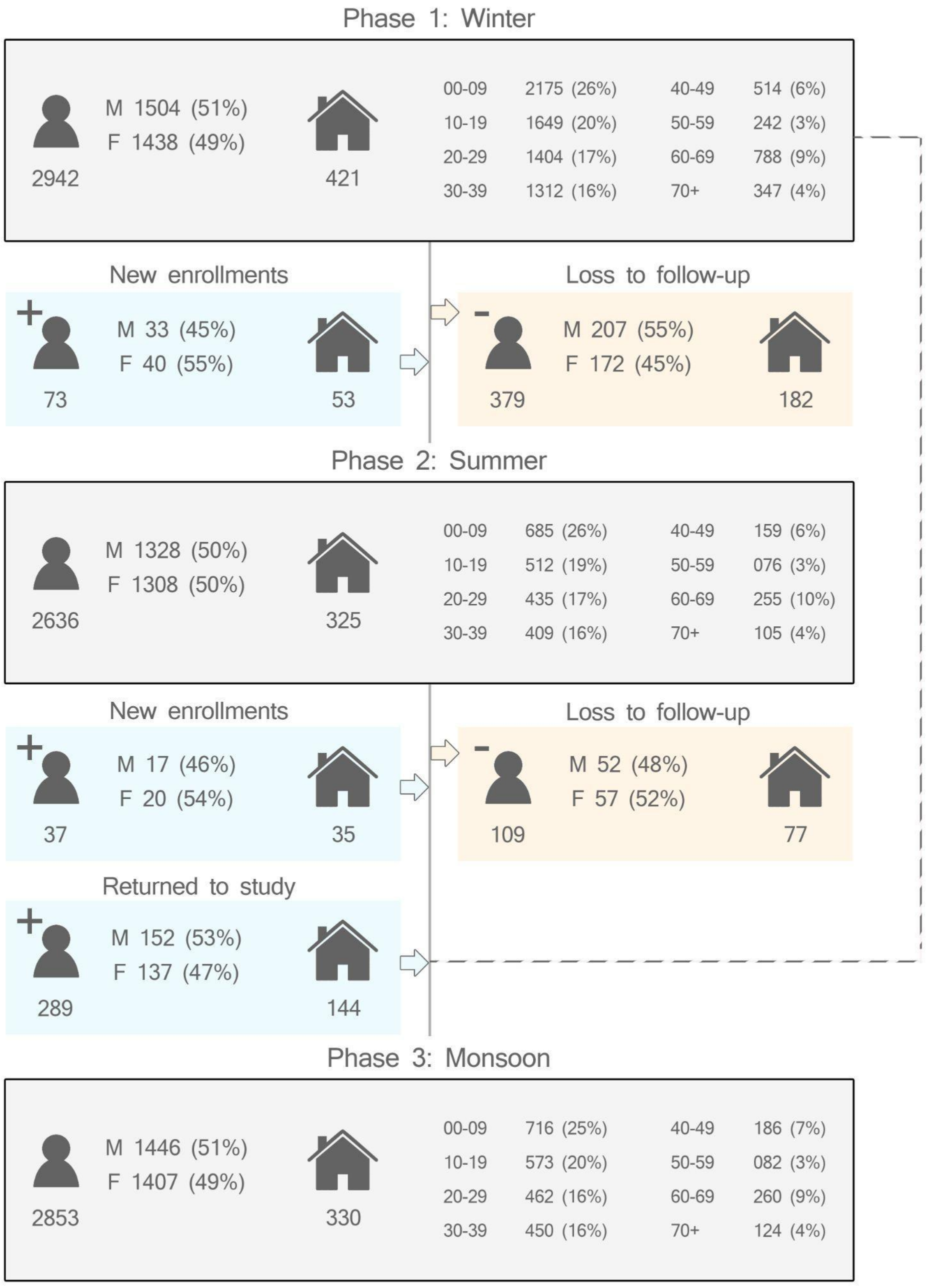


**Fig S1: Respondent characteristics by season.** *The age and gender stratified respondent counts per season are displayed in the grey blocks, while the blue and orange blocks show the loss and gain of respondents in follow-up surveys.*

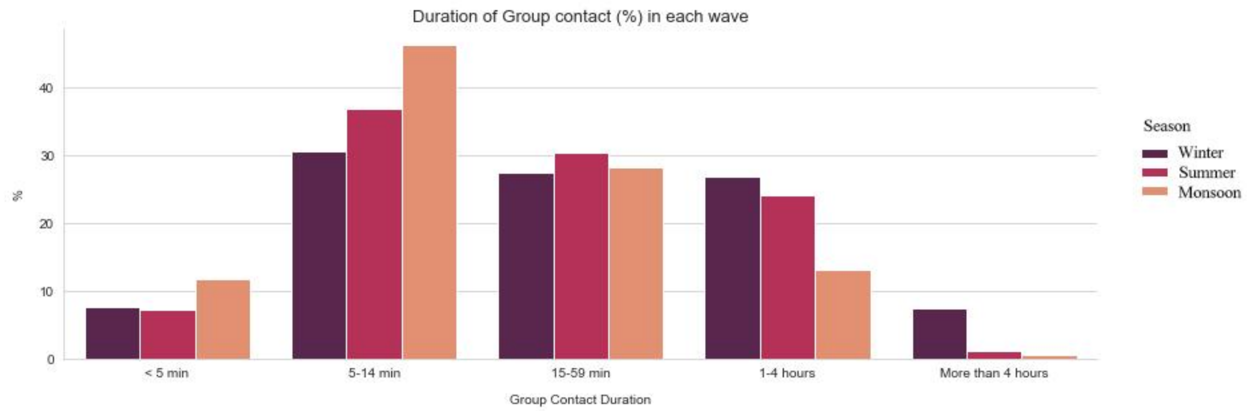


**Fig S2: Percentage of group contacts in a season in each duration bucket.** *Winter had a higher percentage of group contacts that were greater than 4 hours, compared to the other seasons.*

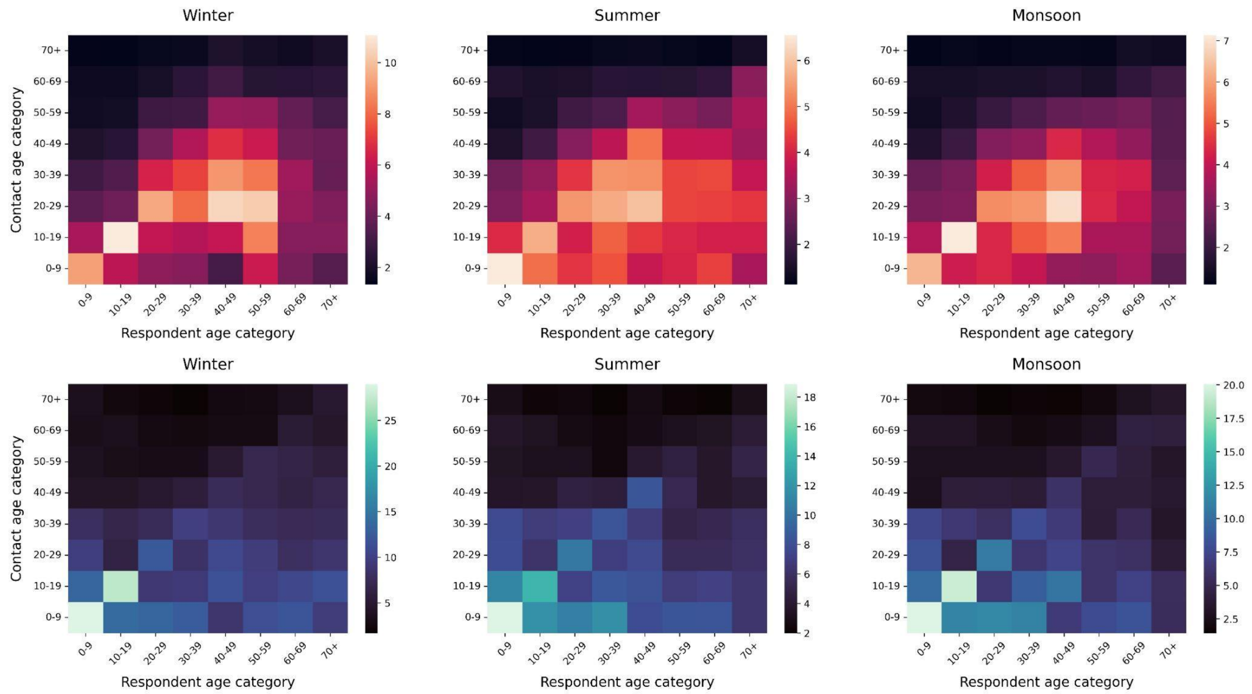


**Fig S3: Number and Durations of Contacts.** *Heatmaps representing mean numbers (red) and durations (blue, in person-hours) of contacts reported between age category dyads. Brighter colours imply a pair of categories with a high number/duration of contacts. Bright diagonal elements suggest some form of age-assortativity.*

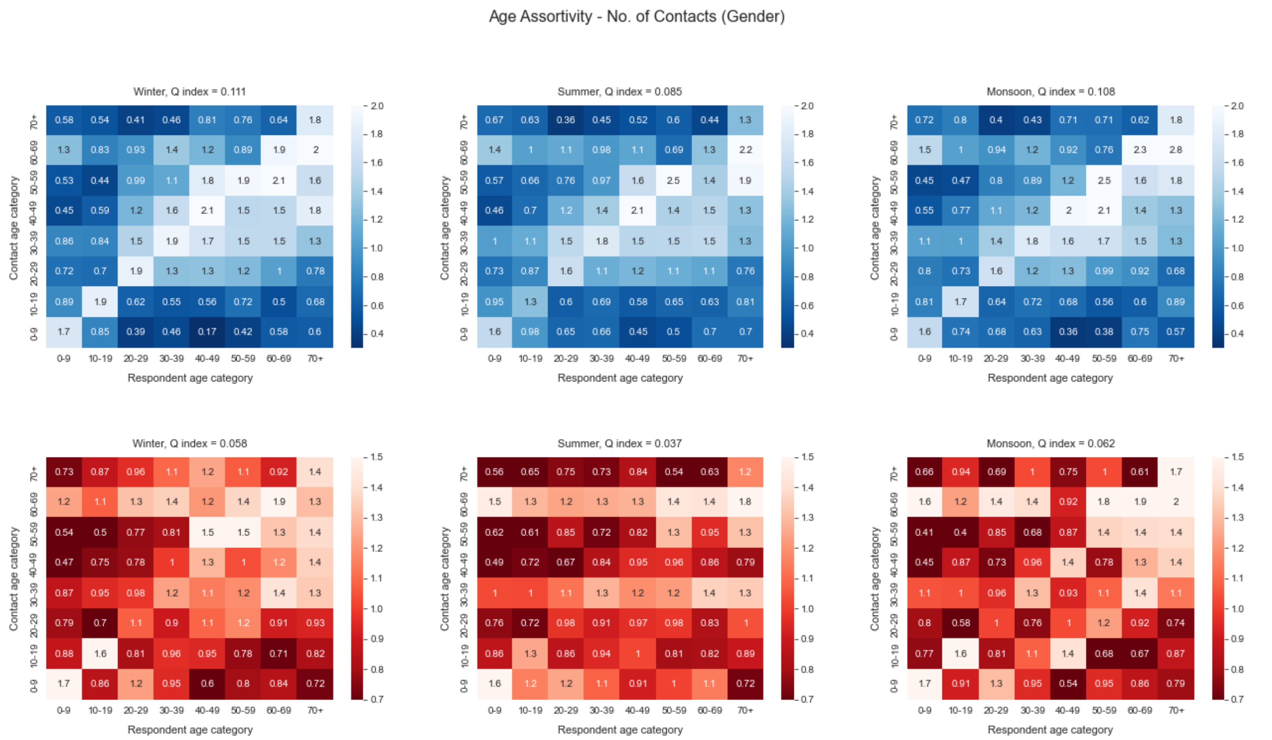


**Fig S4: Number of Contacts by Gender.** *Gender-stratified heat-maps representing the average number of contacts reported between two age categories (Males in blue, females in red). Brighter colours imply a pair of categories with a high number of contacts.*

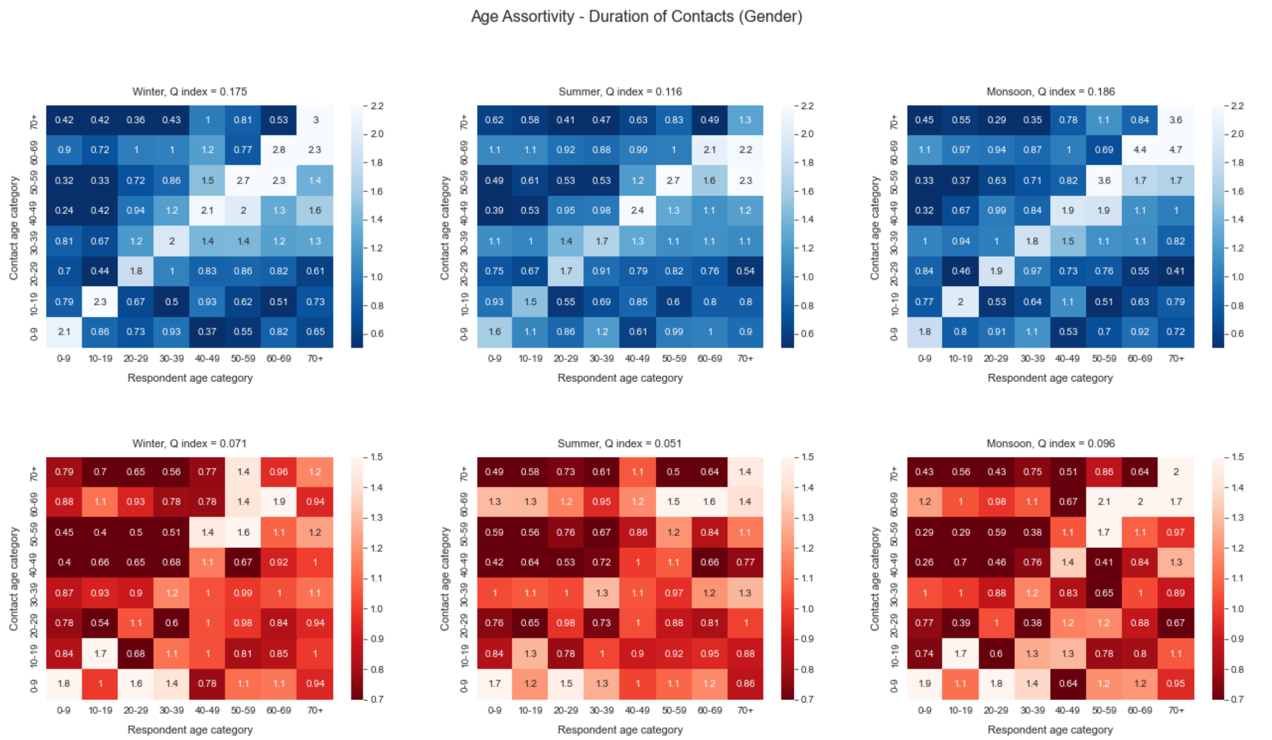

**Fig S5: Duration of Contacts by Gender.** *Gender-stratified matrices representing the average duration of contacts in person-hours reported between two age categories (Males in blue, females in red). Brighter colours imply a pair of categories with a high number of contacts.*

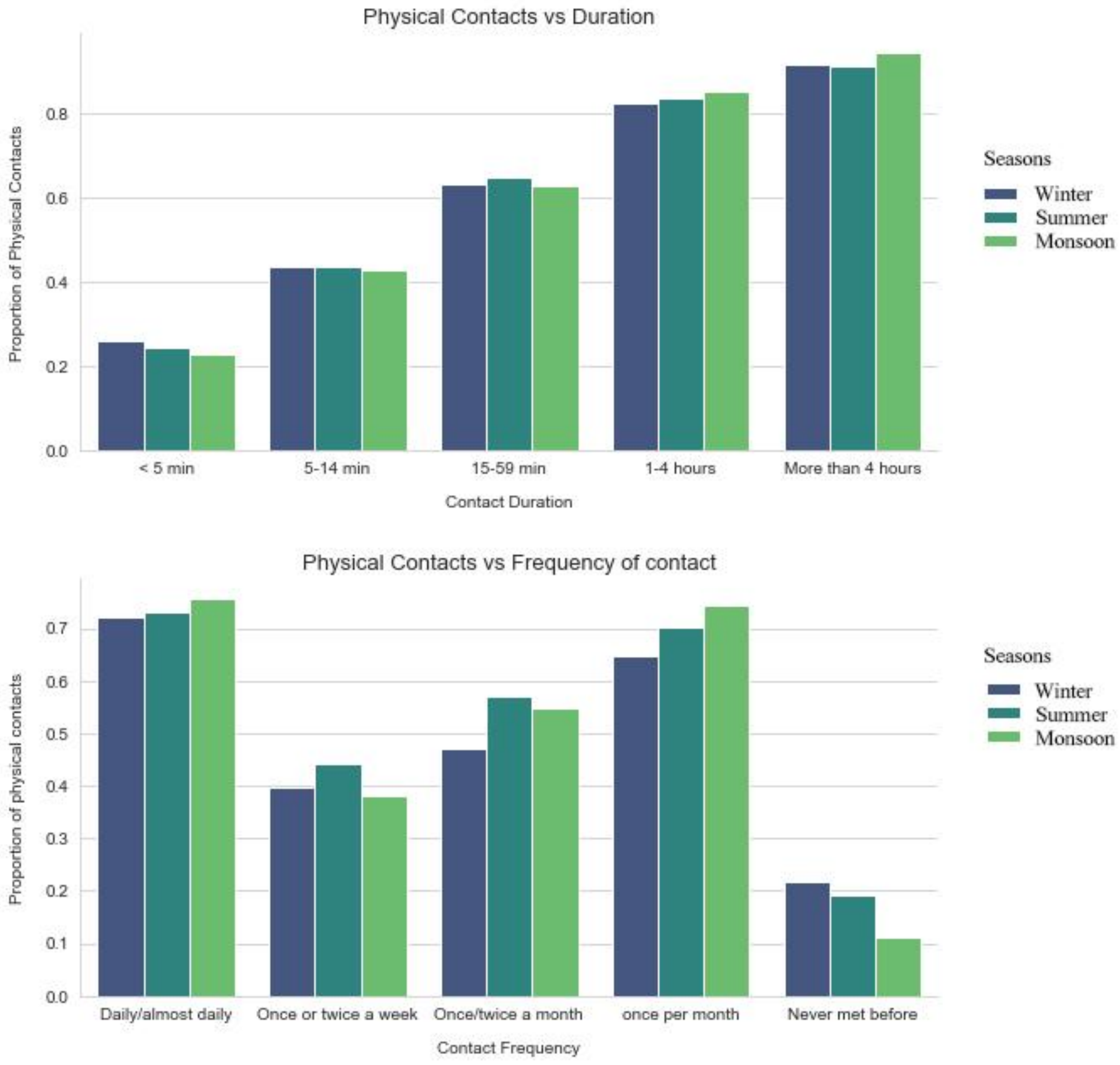


**Fig S6: Proportion of Physical Contacts:** ***(A)*** *Barplots of the proportion of contacts reported to involve physical touch for all three seasons, stratified by the duration of the contact.* ***(B)*** *Proportion of physical contacts for all three seasons stratified by the frequency of which the respondent met the contact*

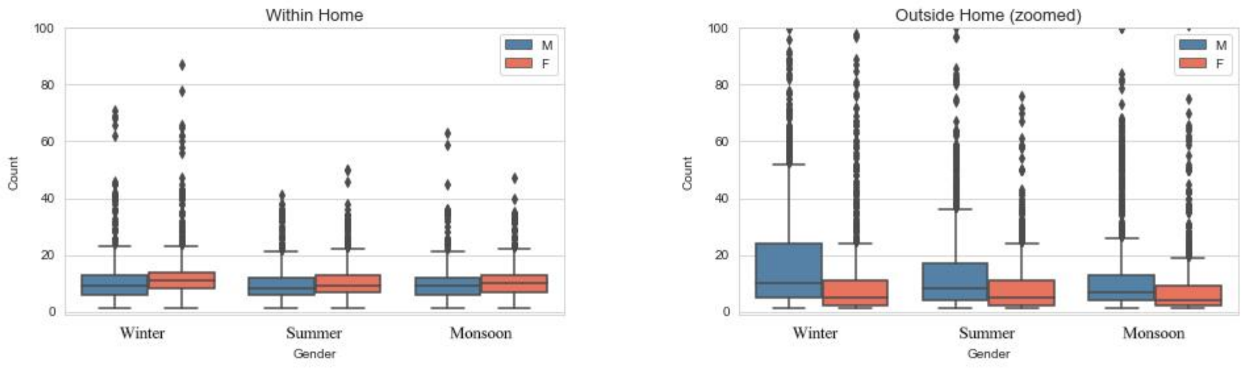
Within Home
Outside Home (zoomed)
Count
Gender
Winter
Summer
Monsoon
M
F

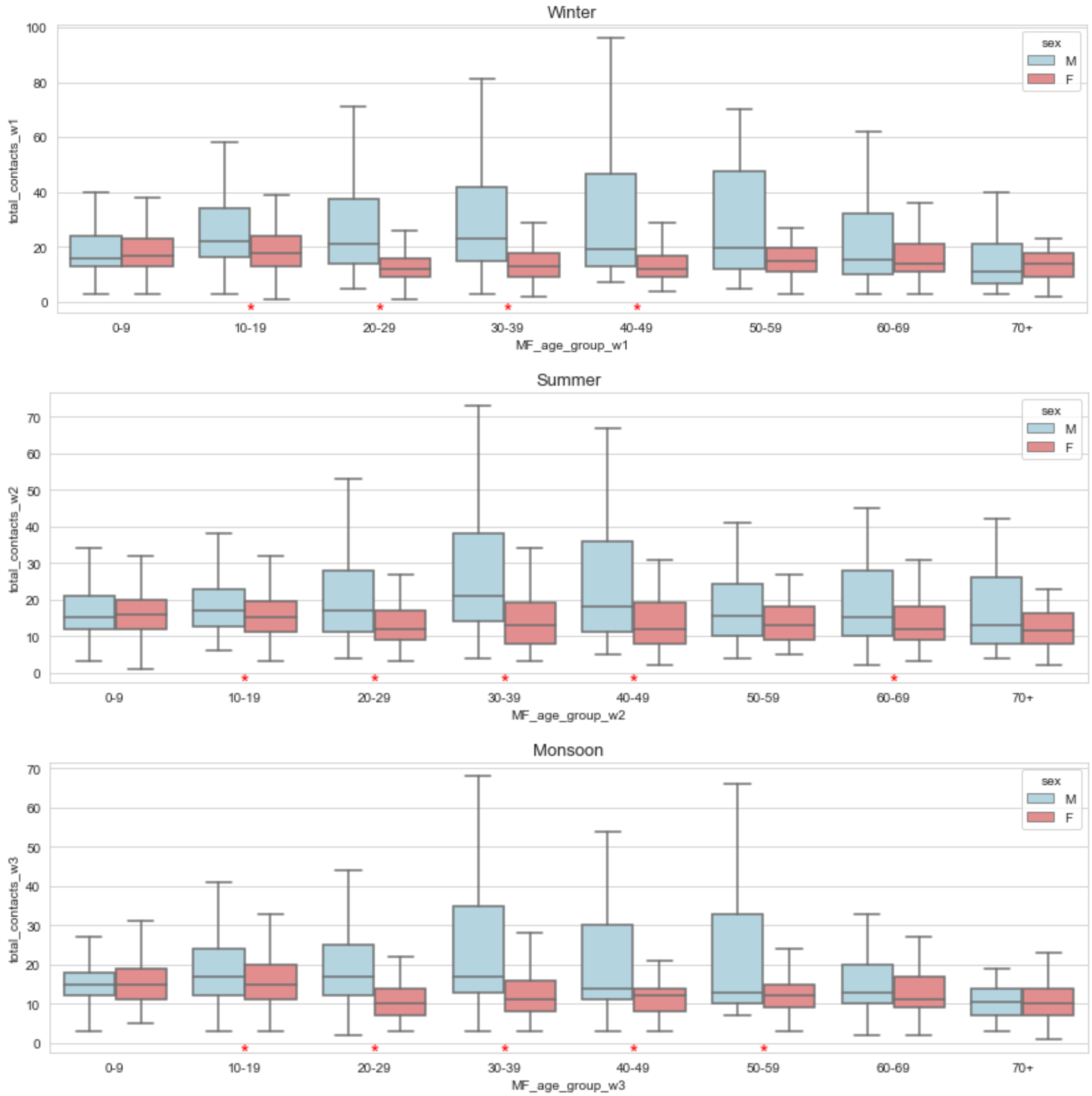
Winter
Summer
Monsoon
total_contacts_w1
total_contacts_w2
total_contacts_w3
MF_age_group_w1
MF_age_group_w2
MF_age_group_w3
sex
M
F
0-9
10-19
20-29
30-39
40-49
50-59
60-69
70+

**Fig S7: Gender Stratified Number of Contacts:** ***(A)*** *Boxplots of the number of contacts that occurred at home reported by males (blue) and females (red).* ***(B)*** *Boxplots of the number of total contacts reported to have occurred outside the home. Note that the upper y limit has been truncated to match that of (A).* ***(C)*** *Boxplots of the number of contacts stratified by gender and age category, across every season.*

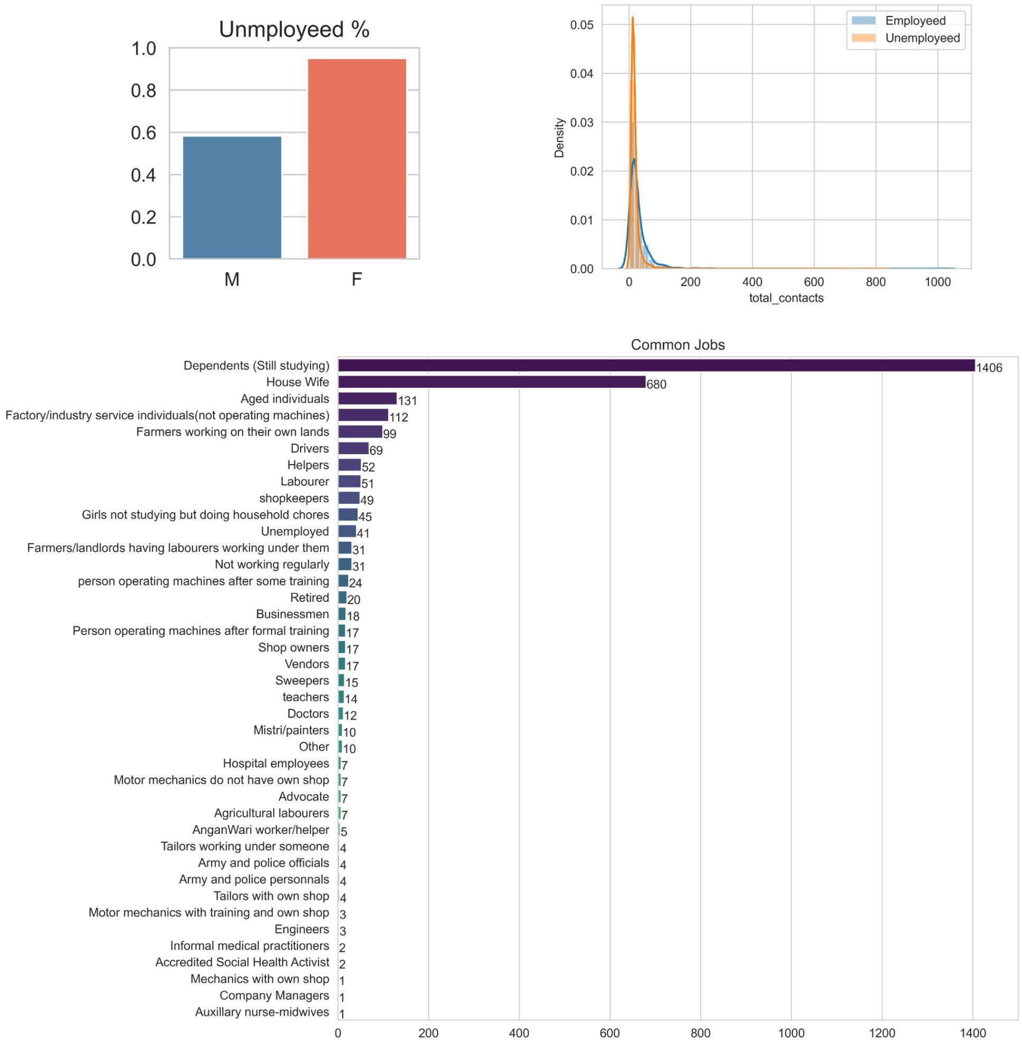


**Fig S8: Occupations and employment outside the home:** ***(A)*** *Percentages of males and females classed as unemployed outside the home.* ***(B)*** *Distribution of total contacts for both employed and unemployed. Solid line represents a gaussian KDE. Note the longer tail on the distribution for employed respondents.* ***(C)*** *Frequencies of occupations (n=3033) among the respondents of the survey.*

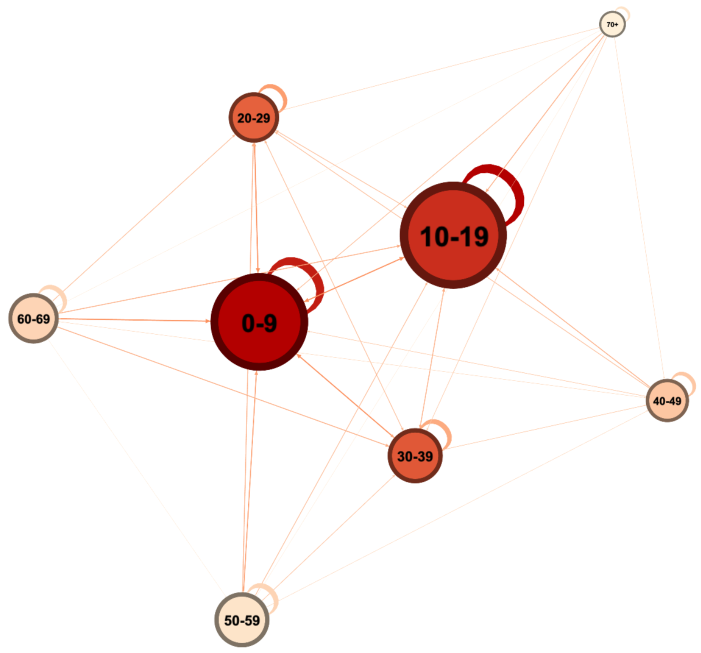


**Fig S9: Social network of total contacts (individual and group) at Ballabgarh.** *Directed graph representing the median number of contacts an age category has with every other age category. Higher numbers are depicted with stronger edge weights. Node sizes represent the total median contacts and node colours represent the PageRank of each node (darker = more important).*

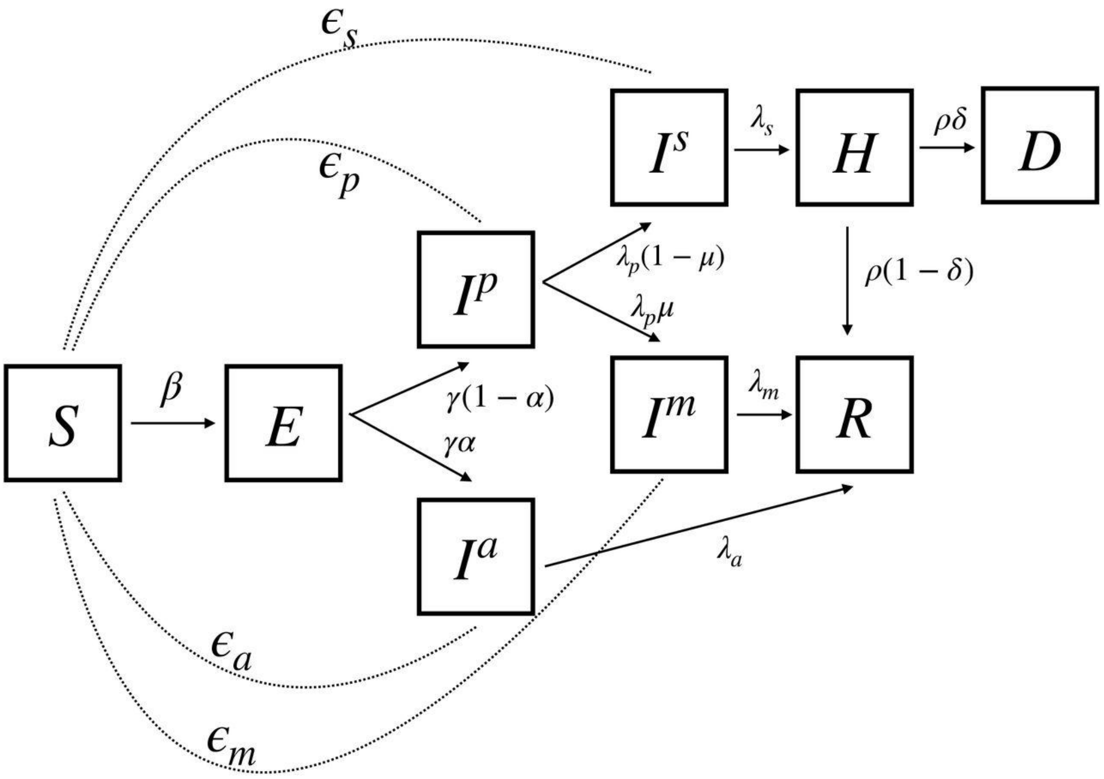

**Fig S10: The compartmental model used in the simulation:** *Figure sourced from Hazra et al.[32] Compartments: S=susceptible, E=exposed,* $I^p$*=Presymptomatic,* $I^a$*=Asymptomatic,* $I^m$*=Mildly Infected,* $I^s$*=Severely Infected, H=Hospitalised, R=Recovered, D=Deceased.*

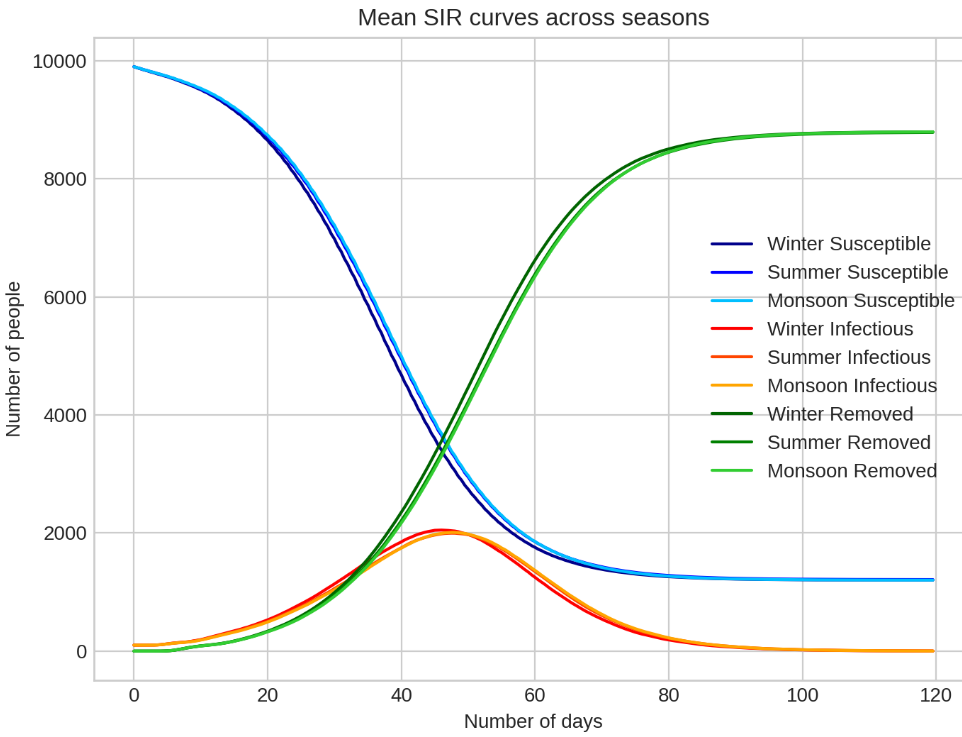


**Fig S11: Mean SIR curves from the simulation.** *Blue, red and green represent susceptible, infectious, and removed counts respectively. Darker colours represent the winter, lighter colours monsoon and summer. Lines are the mean of 50 runs on a population of 10,000 for each season.*

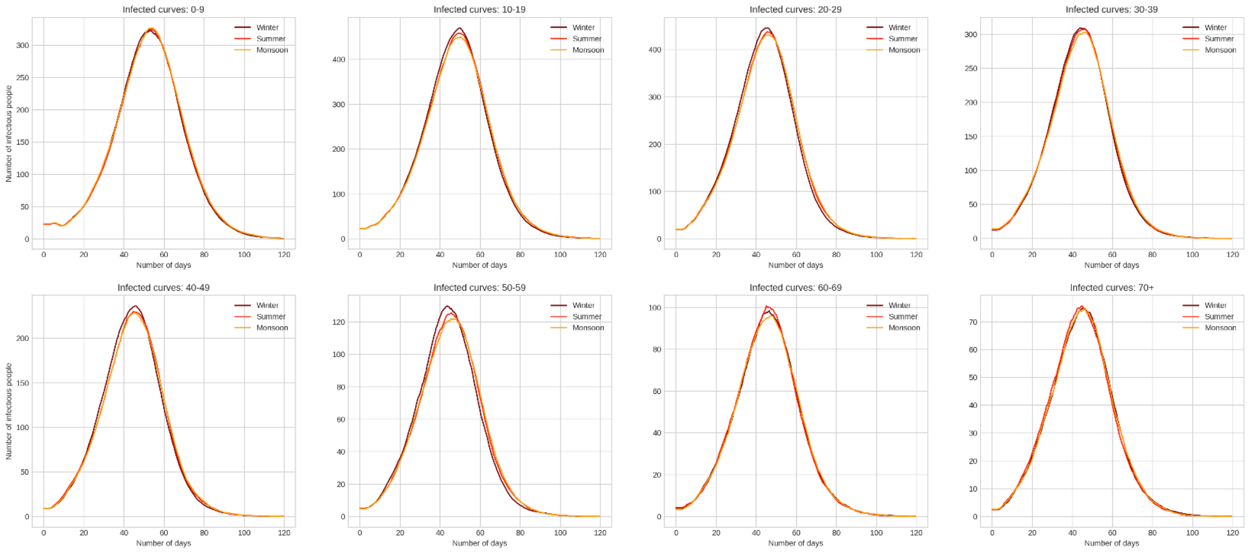


**Fig S12: Age-Stratified infectious curves from the simulation.** *Mean number of infectious people across 50 simulations for every season who belong to a particular age category. Brown, red and orange represent winter, summer and monsoon seasons respectively. Significant difference across seasons is observed in the 50-59 category.*

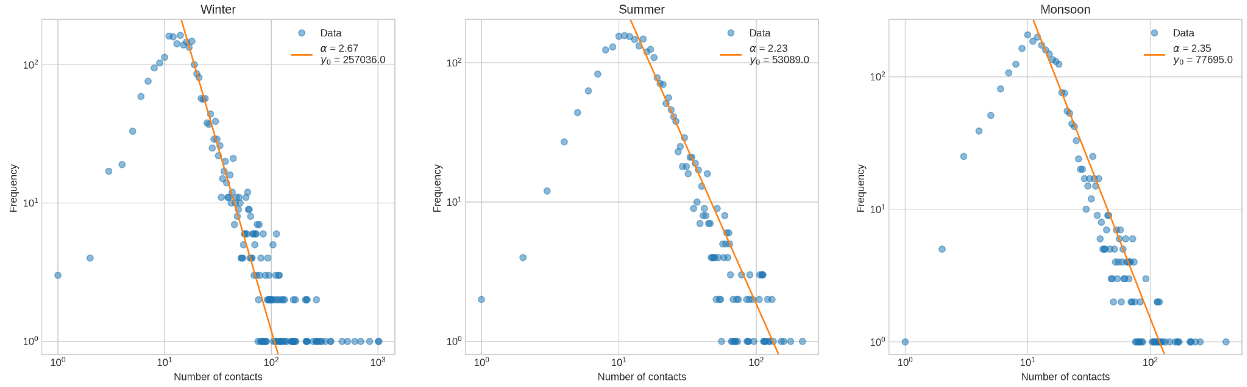


**Fig S13: Distribution of number of contacts.** *The number of individuals who reported a certain amount of total contacts is plotted against the number of total contacts. A curve of* $y = \frac{y_0}{x^\alpha}$ *is fit on the descending section of the data. Note that* $\alpha$ *represents the negative slope of the line on the log-log plot.*

#### Supplementary Tables

| S.No | Attribute | Methodology |
|---|---|---|
| 1 | Dates | All date formats *(31/10/21, 31/10/2021, 31-Oct-21)* were normalised to a common date format. Manual correction of dates was performed based on the season (wave). |
| 2 | Respondent Age | The date of birth of each respondent and the date of interview was used to compute the age. |
| 3 | Contact Gender | Other mentions of the same name were filtered and the mode of the reported genders was used as the gender of the respondent. In case of conflicts, the Genderize API[49] was used to predict the gender based on the respondent name, followed by manual validation. |
| 4 | Group size | Imputed by the median group size of contacts occurring at the same location. For example, for a group contact at home with a missing group size, all group contacts at home were filtered and the median group size was used for imputation. |
| 5 | Duration of group contact | Imputed by the mode duration of group contact occurring at the same location. |
| 6 | Minimum, maximum age in group contact | Imputed by the median minimum and maximum ages of group contacts occurring at the same location. |
| 7 | Contact Age | Contact ages were imputed by sampling from the contact ages of similar respondents. Similar respondents were defined as people who had a similar contact age distribution based on their JS divergence. A Random Forest regression model was trained to predict the JS divergence between two respondents based on features such as their age difference, difference in the number and duration of contacts, proportion of contacts with males and more. A set of similar respondents was obtained for a given respondent and the missing contact ages were sampled from the contact age distribution of these respondents. |

**Table S1. Data cleaning and imputation methodology.**

| S.No | Group contact category | Keywords |
|---|---|---|
| 1 | Chatting | chating, chatting, chhatin |
| 2 | Death | condolence, death, derth, dearth, shok, sok |
| 3 | Festival | birthday, festival, holi, kanjikey, namkaran, raksha, rasam, sakrant |
| 4 | Game | cricket, game, kabbadi, play, sports |
| 5 | Madarsa | madarsa, madrasa, madatsra, madatrsa |
| 6 | Politics | chunav, election, rally, rss, vote, voter |
| 7 | School | aanganwari, aganwadi, anganwadi, anganwari, class, coaching, college, colledge, school, taleem, talim, tuition, tution |
| 8 | Shop | buy, dairy, dukan, market, milk, purchase, shoop, shop, vendor |
| 9 | Transport | auto, bus, ola, taxi, train, transport, travel |
| 10 | Wedding | barat, baraat, function, fuction, kanya, marriage, ring, sagai, sangeet, shadi, wedding |
| 11 | Work | business, company, duty, labour, office, job, polymed, wager, work |
| 12 | Worship | bhagvat, bhagwat, bhagwath, eid, jagran, jagrata, kirtan, langar, mandir, masjid, maszid, mosk, mosque, namaj, pray, ramayan, satsang, satssang, temple, worship |

**Table S2. Keywords to map group contact reasons (free-write strings) to predefined categories.**

| Month | chatting | death | festival | game | madarsa | other | politics | school | shop | transport | wedding | work | worship |
|---|---|---|---|---|---|---|---|---|---|---|---|---|---|
| Jan | 290 | 255 | 270 | 119 | 744 | 833 | 4638 | 370 | 520 | 63 | 708 | 2458 | 110 |
| Feb | 123 | 150 | 0 | 85 | 185 | 556 | 0 | 606 | 987 | 665 | 2023 | 1735 | 95 |
| Mar | 77 | 215 | 216 | 97 | 100 | 397 | 0 | 358 | 839 | 0 | 294 | 160 | 192 |
| Apr | 174 | 140 | 0 | 82 | 40 | 605 | 0 | 110 | 183 | 28 | 625 | 506 | 167 |
| May | 110 | 180 | 25 | 158 | 135 | 490 | 0 | 354 | 575 | 258 | 422 | 1199 | 98 |
| Jun | 43 | 85 | 30 | 27 | 0 | 393 | 0 | 28 | 1012 | 315 | 20 | 2326 | 10 |
| Jul | 35 | 0 | 20 | 10 | 15 | 94 | 0 | 125 | 415 | 0 | 98 | 480 | 105 |
| Aug | 40 | 355 | 180 | 50 | 0 | 375 | 0 | 483 | 217 | 100 | 60 | 814 | 60 |
| Sep | 230 | 50 | 40 | 199 | 181 | 278 | 0 | 82 | 440 | 30 | 50 | 1123 | 153 |
| Oct | 190 | 350 | 50 | 130 | 70 | 1070 | 0 | 1064 | 1466 | 170 | 355 | 3716 | 285 |
| Nov | 90 | 100 | 30 | 131 | 0 | 1509 | 30 | 1108 | 1615 | 98 | 2200 | 354 | 688 |
| Dec | 90 | 312 | 0 | 7 | 350 | 1600 | 80 | 385 | 865 | 145 | 722 | 343 | 235 |

**Table S3. Month and location wise break down of the number of people met in group contacts**

| Parameter | Description | Distribution (mean, sd) |
|---|---|---|
| *t_exposed* | Length of time after exposure before individual is infectious | lognormal (4.5, 1.5)[50] |
| *t_presymp* | Length of time after individual is infectious until symptoms occur | lognormal (1.1, 0.9)[51],[50] |
| *t_asymp* | Length of time from infectiousness onset to recovery for asymptomatic cases | lognormal (8.0, 2.0)[52] |
| *t_mild* | Length of time a person spends in the mildly infected compartment (infectiousness onset to recovery/severity) | lognormal (8.0, 2.0)[52] |
| *t_severe* | Length of time from the occurrence of severe symptoms to the person needing hospitalisation | lognormal (1.5, 2.0)[53],[54] |
| *t_hospitalized* | Length of time a person spends in the hospitalised compartment | lognormal (18.1, 6.3)[55] |

**Table S4. Rates and parameters used in the agent-based simulation:** *The rates used are extracted from the Covasim model developed by Kerr et al.[31]*

| | 0-9 | 10-19 | 20-29 | 30-39 | 40-49 | 50-59 | 60-69 | 70-79 | 80-89 | 90+ |
|---|---|---|---|---|---|---|---|---|---|---|
| *r_sus[9]* | 0.34 | 0.67 | 1.00 | 1.00 | 1.00 | 1.00 | 1.00 | 1.24 | 1.47 | 1.47 |
| *p_symp [55,56]* | 0.50 | 0.55 | 0.60 | 0.65 | 0.70 | 0.75 | 0.80 | 0.85 | 0.90 | 0.90 |
| *p_severe[55,56]* | 0.005 | 0.00165 | 0.00720 | 0.02080 | 0.03430 | 0.07650 | 0.13280 | 0.20655 | 0.24570 | 0.24570 |
| *p_deceased[57],[58]* | 0.00002 | 0.00002 | 0.00010 | 0.00032 | 0.00098 | 0.00265 | 0.00766 | 0.02439 | 0.08292 | 0.16190 |

**Table S5. Age Stratified rates used in the simulation:** *r_sus is relative susceptibility, and is multiplied to the value of a target's beta. p_symp represents the probability of showing symptoms, p_severe the probability of displaying severe symptoms given the person is infected, and p_deceased the probability of death given the person has severe symptoms. Rates taken from the Covasim model developed by Kerr et al.[31]*

**References**

### References


1. Hazra, D. K. *et al.* The INDSCI-SIM model for COVID-19 in India. *medRxiv* 2021.06.02.21258203 (2021).
2. Genderize.io. https://genderize.io/.
3. Lauer, S. A. *et al.* The Incubation Period of Coronavirus Disease 2019 (COVID-19) From Publicly Reported Confirmed Cases: Estimation and Application. *Ann. Intern. Med.* **172**, (2020).
4. Linton, N. M. *et al.* Incubation Period and Other Epidemiological Characteristics of 2019 Novel Coronavirus Infections with Right Truncation: A Statistical Analysis of Publicly Available Case Data. *J. Clin. Med. Res.* **9**, 538 (2020).
5. Wölfel, R. *et al.* Virological assessment of hospitalized patients with COVID-2019. *Nature* **581**, 465–469 (2020).
6. Chen, J. *et al.* Clinical progression of patients with COVID-19 in Shanghai, China. *J. Infect.* **80**,

(2020).

7. Wang, D. *et al.* Clinical Characteristics of 138 Hospitalized Patients With 2019 Novel Coronavirus–Infected Pneumonia in Wuhan, China. *JAMA* **323**, 1061–1069 (2020).
8. Estimates of the severity of coronavirus disease 2019: a model-based analysis. *Lancet Infect. Dis.* **20**, 669–677 (2020).
9. Kerr, C. C. *et al.* Covasim: An agent-based model of COVID-19 dynamics and interventions. *PLoS Comput. Biol.* **17**, e1009149 (2021).
10. Zhang, J. *et al.* Changes in contact patterns shape the dynamics of the COVID-19 outbreak in China. *Science* **368**, (2020).
11. Report 9 - Impact of non-pharmaceutical interventions (NPIs) to reduce COVID-19 mortality and healthcare demand. *Imperial College London* http://www.imperial.ac.uk/medicine/departments/school-public-health/infectious-disease-epidemiology/mrc-global-infectious-disease-analysis/covid-19/report-9-impact-of-npis-on-covid-19/.
12. O'Driscoll, M. *et al.* Age-specific mortality and immunity patterns of SARS-CoV-2. *Nature* **590**, (2021).
13. Report 34 - COVID-19 Infection Fatality Ratio Estimates from Seroprevalence. *Imperial College London* http://www.imperial.ac.uk/medicine/departments/school-public-health/infectious-disease-epidemiology/mrc-global-infectious-disease-analysis/covid-19/report-34-ifr/.
